# Clinical Application of Large Language Models for Breast Conditions: A Systematic Review

**DOI:** 10.1101/2024.08.31.24312542

**Authors:** Billy Ho Hung Cheung, Karen Gwyn Poon, Cheuk Fai Lai, Ka Chun Lam, Michael Co, Ava Kwong

**Author notes:** **Corresponding author:** Professor Ava Kwong, Daniel C K Yu Professor in Breast Cancer Research, Clinical Professor, Chief of Division of Breast Surgery, Department of Surgery, Li Ka Shing Faculty of Medicine, The University of Hong Kong, Queen Mary Hospital, Hong Kong SAR, China (; Email address).

## Abstract

**Background:** The application of artificial intelligence (AI) like Large Language Models (LLM) into the healthcare system has been a frequently discussed topic in recent years.

**Materials and Methods:** We conducted a systemic review on primary studies about the applications of LLM in breast conditions. The studies are then categorized into their respective domains, namely diagnosis, management recommendations and communication for patients.

**Results:** The diagnostic accuracy ranged from 74.3% to 99.6% across different investigation modalities. The concordance of management recommendations ranged from 50% to 70% while the prognostic evaluation of breast cancer patients of distant recurrence showed an accuracy of 75% to 88%. In regards to patient communication, it is revealed that 18-30% of the references used by the LLM were irrelevant.

**Conclusion:** This study highlights the potential benefits of LLM in strengthening patient communication, diagnose and management of patients with breast conditions. With standardized protocol and guideline to minimize potential risks, LLM can be a valuable tool to support future clinicians in the field of breast management.

## 1. Introduction

Artificial intelligence (AI) has shown a promising future for enhancing various aspects of healthcare, from diagnosis to treatment planning to patient education. [1] Large language models (LLMs), a type of AI system that can interpret and generate human language, have gained particular interest for their potential in assisting with clinical tasks which involve unstructured text data. [2] LLMs, like ChatGPT, have demonstrated impressive performance on general knowledge and language understanding benchmarks. [3] However, their utility and limitations for specific clinical applications remain to be established.

Breast cancer is one of the most common malignancies worldwide with an annual incidence of 2.3 million.[4] Breast conditions-related clinical visits make up around three percent of primary health care visits among women. [5] They range from benign diseases and breast cancer with a wide variety of management. However, due to the overloading number of patients, the waiting time for patients’ clinical assessment can be significantly prolonged. [6] Moreover, the clinical evaluation for breast related diseases, which involves triple assessment and personalised approach, can be strained due to time restraints and limited resources. [6]

Hence, it is our belief that the management of patients with breast conditions can be an important area where AI like LLMs could potentially aid clinicians and patients. Accurate interpretation of imaging and pathology reports is crucial for breast diagnosis,[7] while personalized management requires complex decision-making based on multiple clinical factors. Effective communication of information to patients is also key for shared decision-making and treatment compliance. [8] Prior studies have explored applications of narrow AI systems for breast imaging interpretation and clinical decision support. [9, 10] However, the use of more flexible and comprehensive LLMs for breast conditions has not been systematically examined.

The objective of this systematic review is to evaluate the current evidence on the application of LLMs in breast conditions. We further categorise them into diagnosis, management, and patient communication. Specifically, we aim to address three key questions: 1) What is the diagnostic accuracy of LLMs for interpreting breast imaging and pathology reports? 2) How well do LLM management recommendations align with those of multidisciplinary teams? 3) What is the quality and limitations of LLM-generated information for patients on breast health topics? By critically appraising and synthesizing the available studies, we seek to identify promising use cases, current gaps, and future directions for the development and validation of LLMs in breast care.

## 2. Materials and Methods

We conducted a systematic search of PubMed, SCOPUS, and Google Scholar databases for studies published up to December 31, 2023 according to the systematic review using the Preferred Reporting Items for Systematic Reviews and Meta-Analyses (PRISMA) guidelines. [11] The search query included terms related to large language models “ChatGPT” OR “BARD” OR “LLAMA” OR “Large language model” OR “LAMDA” OR “GPT” OR “GPT” OR “natural language model” OR”natural language process*”) in combination with “breast”.

We included only primary studies that assessed the performance or application of LLMs for any of the following in relation to breast conditions. These studies were then categorised according to the predefined domains, including diagnosis, management recommendations, and communication for patients. Commentaries, editorials, and reviews were excluded. Non-LLM based narrow AI systems were also excluded. Two reviewers independently screened titles and abstracts, followed by full-text review of potentially eligible studies. Disagreements were resolved by consensus or consultation with a third reviewer.

## 3. Theory/Calculation

For each included study, we extracted information on the study design, data sources, the LLM studied, comparison method, sample size, and main outcomes. For diagnostic studies, we extracted information regarding the metrics of accuracy, such as sensitivity, specificity, and area under the receiver operating curve (AUC). For management studies, we extracted concordance rates between LLM and human expert recommendations. For patient communication studies, we summarized the quality assessments and any limitations that are identified.

We synthesized results narratively and in tables, stratified by application area (diagnosis, management, patient communication). Meta-analysis was not proceeded due to the limited data.

## 4. Results

### 4.1. Study Characteristics

The search yielded 173 records, of which 17 studies met inclusion criteria (Figure 1). Eight studies evaluated LLMs for breast diagnosis,[12-19] three for management,[20-22] two for prognosis [23, 24] and four for patient communication. [25-28] Two studies assessed multiple applications. [26, 27] The most commonly used LLM was ChatGPT (n=6), followed by custom models based on BERT or GPT architectures. Most studies were conducted in the USA or Europe.

**Figure 1:**
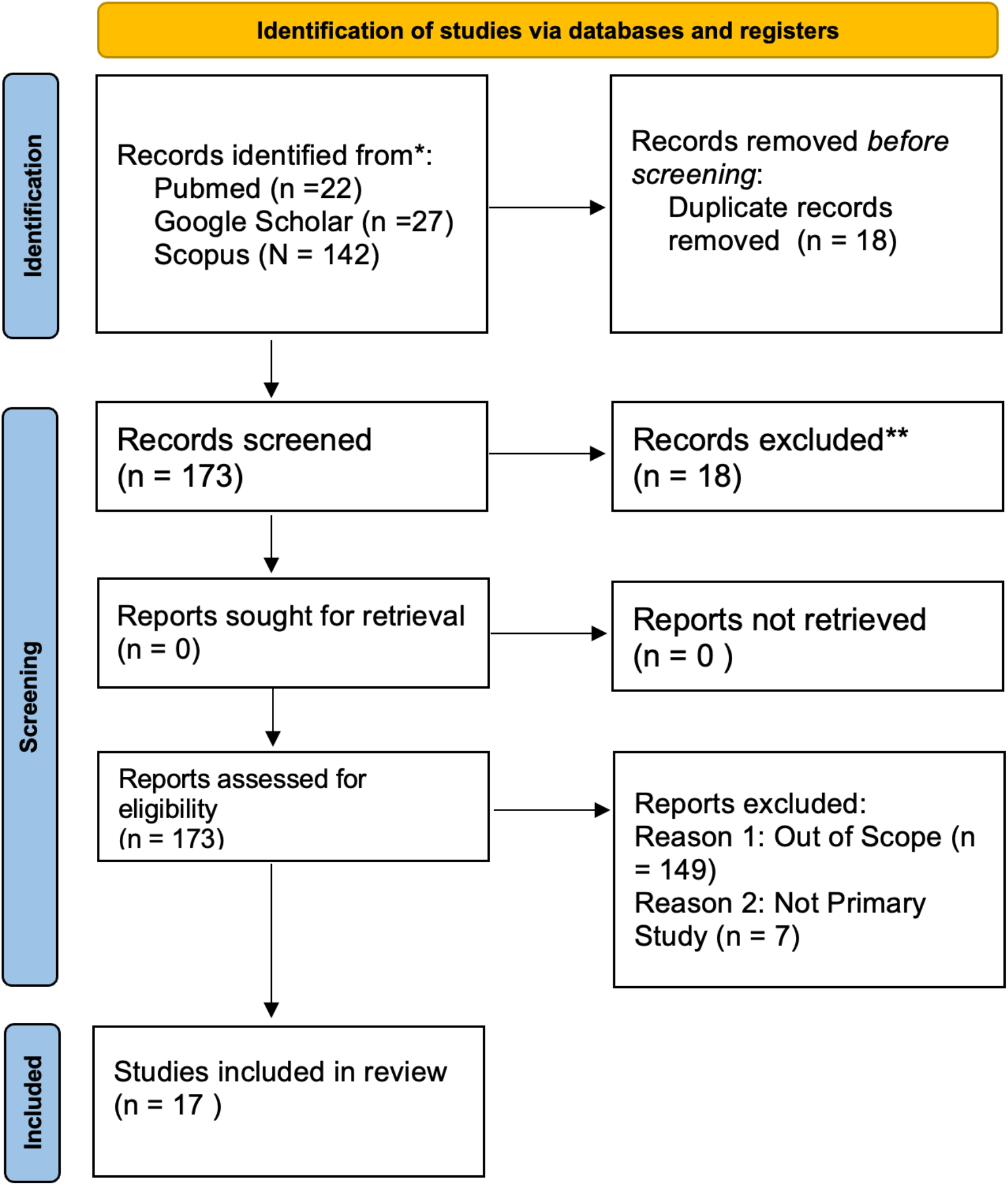
Systematic review using the PRISMA guideline

### 4.2. Diagnostic Accuracy

Seven studies use developed AI models to diagnose patients with radiological free text reports, ranging between 300 to 79312 reports from each study. The accuracy of LLMs for classifying breast conditions based on breast imaging reports (mammograms, ultrasound, MRI) and pathology reports (Table 1). Accuracies ranged from 74.3% to 99.6% across modalities, with the highest precision for MRI-based lesion detection (99.6%) and lowest for BI-RADS category assignment on mammogram-ultrasound (74.3%). [17] Two studies found that LLM accuracies were comparable to manual interpretation by radiologists. [13, 15]

**Table 1.** Summary table for diagnostic report usage.

| Imaging modalities | No. of papers |
| --- | --- |
| Mammogram | 2 |
| Ultrasound and Mammogram | 1 |
| MRI | 2 |
| Pathological Report | 1 |
| Ultrasound and Pathological Report | 1 |

Only one study evaluated a publicly-available LLM (ChatGPT) as an aid for radiologists, finding acceptable accuracy (83%) but noting limitations in currency of knowledge and potential for incorrect or fraudulent output. [18] Other studies used custom-developed LLMs based on transformer architectures and trained on site-specific imaging reports. Common challenges included extraction of granular radiographic features, assignment to proper BI-RADS categories, and integration into clinical workflows.

### 4.3. Management Recommendations and Prognosis

Three studies compared management recommendations of LLMs to those of multidisciplinary tumor boards (MDTs) for breast cancer cases, with population sizes ranging from 10 to 20 patients (Table 2). Overall, the concordance ranged from 50% (for cases including benign/precancerous lesions) to 70% (for invasive cancers only). [21, 22] Concordance rate was highest for recommendations on radiotherapy (95%), followed by chemotherapy (94.7%), endocrine therapy (75%), genetic testing (70%). [20] Discordant recommendations were often due to LLMs not considering patient age, performance status, or specific tumor features. In addition, LLMs sometimes proposed fraudulent decisions, such as recommending extensive genetic testing for patients without positive family history of breast cancer and advising against re-excision for cases with positive surgical resection margins. Over-treatment with chemotherapy and under-treatment with surgery were also observed. The authors noted that LLMs currently have limited suitability as a decision aid for breast cancer management due to medicolegal issues and potential for harm from erroneous recommendations.

**Table 2.**
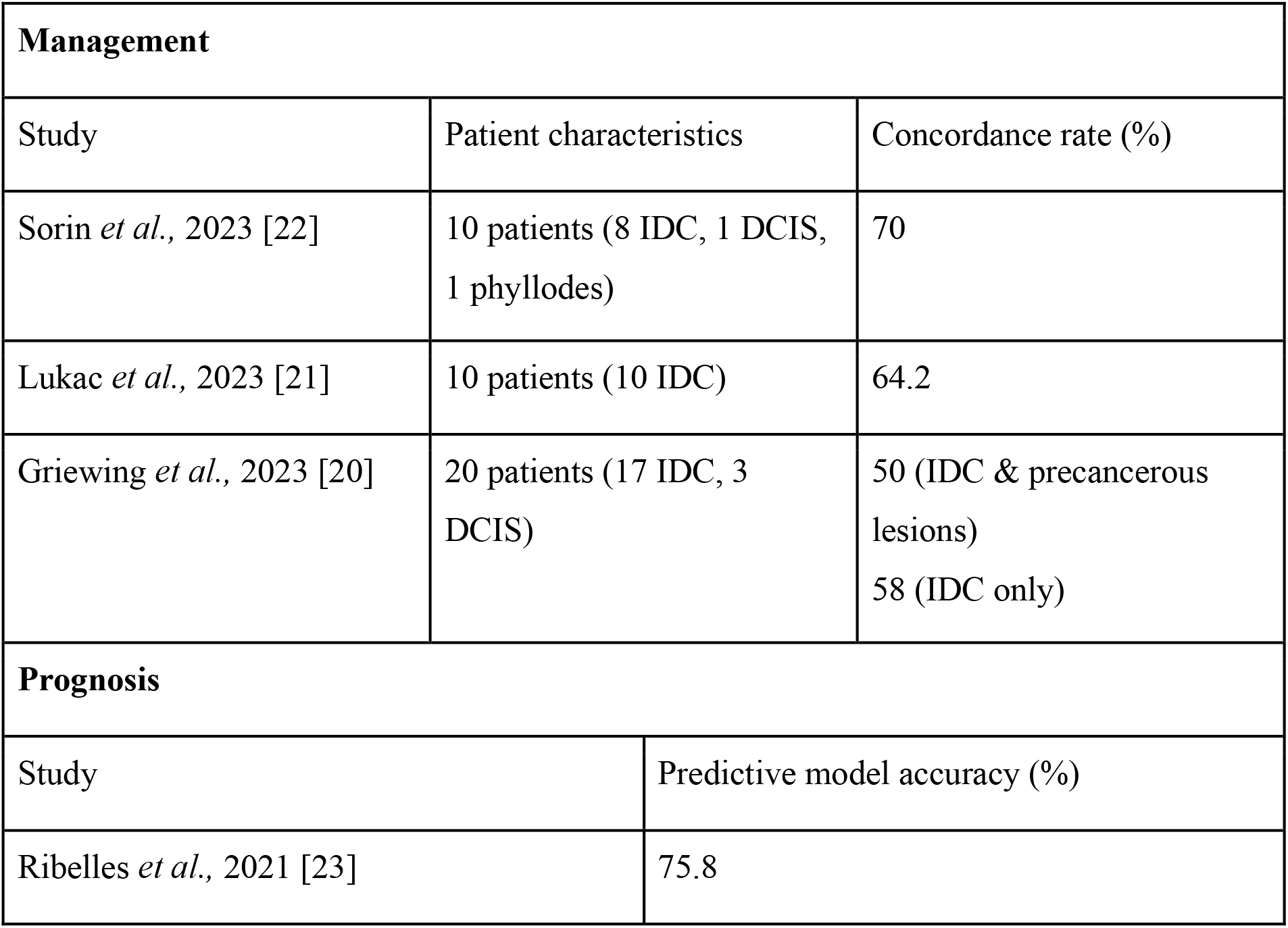
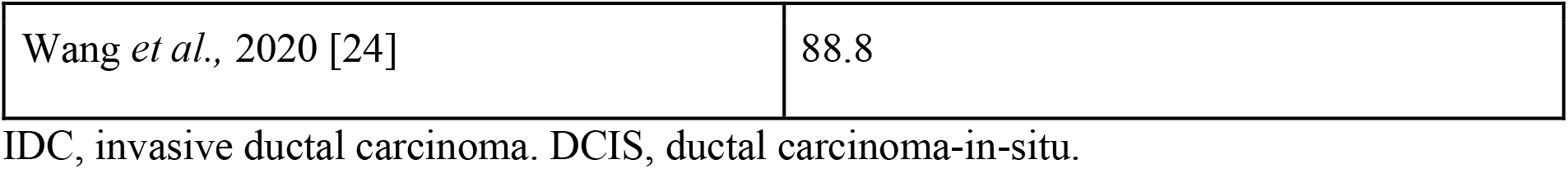
Summary table on management and prognosis.

| Management |  |  |
| --- | --- | --- |
| Study | Patient characteristics | Concordance rate (%) |
| Sorin *et al.*, 2023 [22] | 10 patients (8 IDC, 1 DCIS, 1 phyllodes) | 70 |
| Lukac *et al.*, 2023 [21] | 10 patients (10 IDC) | 64.2 |
| Griewing *et al.*, 2023 [20] | 20 patients (17 IDC, 3 DCIS) | 50 (IDC & precancerous lesions)<br>58 (IDC only) |
| Prognosis |  |  |
| Study | Predictive model accuracy (%) |  |
| Ribelles *et al.*, 2021 [23] | 75.8 |  |

|  |  |
| --- | --- |
| Wang *et al.*, 2020 [24] | 88.8 |

IDC, invasive ductal carcinoma. DCIS, ductal carcinoma-in-situ.

Furthermore, two studies demonstrated the application of LLM-based predictive models on real breast cancer patients for prognostic evaluation, which showed an accuracy of 75% to 88% in the prediction of distant breast cancer recurrence. [23, 24]

### 4.4. Patient Communication

Four studies assessed the quality of information produced by LLMs in response to common patient questions about breast health topics (Table 3). All used ChatGPT, either version 3.5 or Number of questions included in each study varies, ranging from 6 to 25, each study recruited 3-5 experienced breast surgeons/ specialists to comment on the answers generated by LLMs. Across studies, clinician quality ratings of LLM-generated content averaged 4.2 on a 5-point scale, indicating high overall quality. LLMs were noted to provide comprehensive, technically accurate, and well-organized answers, generally outperforming search engines. [26, 28]

**Table 3.**
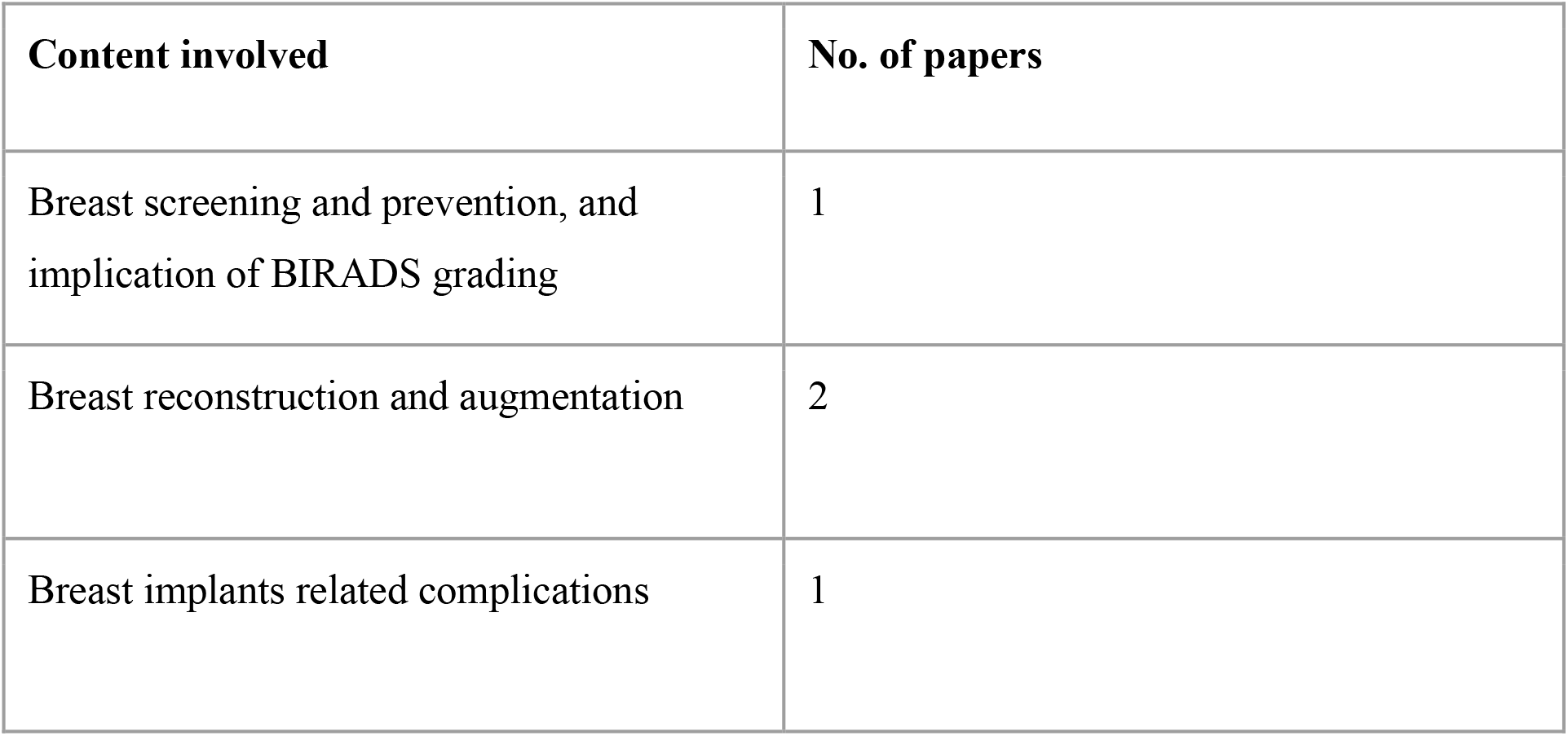
Summary table on patient communication.

However, several important limitations were identified. Across studies, 18-30% of references cited by ChatGPT were inaccessible, irrelevant, or fabricated, raising concerns about information reliability. [25-27] ChatGPT sometimes gave outdated information, such as not reflecting the latest FDA regulations on breast implants. [28] It also tended to provide generic rather than patient-specific recommendations. [28] Additionally, ChatGPT responses were found to vary based on prompt wordings, occasionally introducing inaccurate or biased content [25]. Google searches were noted to provide more reliable and up-to-date references, especially for controversial topics, namely breast implant illness. [26, 27]

## 5. Discussion

This systematic review shows that LLMs such as ChatGPT have the potential in making clinical diagnosis, formulating individualised management plans, generating prognostic prediction and acting as a patient communication tool for health enquiries. However, significant limitations and potential risks were also identified, highlighting the need for further validation and refinement before clinical deployment.

In the diagnostic domain, LLMs achieved high accuracy for extracting key findings and assigning BI-RADS categories across mammography, ultrasound, MRI, and pathology reports. The performance was often comparable to expert human reviewers, supporting the potential for LLMs to enhance the efficiency and consistency of report interpretation. However, most studies used custom-developed LLMs trained on site-specific data, thus limiting generalizability. One study that evaluates a general-purpose LLM (ChatGPT), found lower sensitivity when compared to usual care for screening mammograms. [18] This underscores the importance of training LLMs on large, diverse datasets reflective of real-world practice. Future work should also establish benchmarks and best practices for assessing LLM performance, as metrics varied widely across studies.

For management, LLMs showed moderate concordance (50-70%) with multidisciplinary tumor board decisions for breast cancer cases. Agreement was highest for recommending standard systemic therapies but lower for nuanced surgical and genetic testing decisions. Importantly, LLMs sometimes gave dangerous recommendations, such as advising against re-excision for positive margins or recommending excessive genetic testing.[20] Over- and under-treatment were also observed relative to guideline-concordant care. These findings caution against using LLMs as autonomous decision-making tools, as they lack the clinical judgment to weigh competing factors and may not reflect the latest evidence. Instead, LLMs may be best positioned as aids to prompt consideration of management options, with final decisions made by human experts. Rigorous testing in prospective studies and refinement on large, curated oncology datasets will be necessary before considering deployment.

In the realm of patient communication, LLMs like ChatGPT generated coherent, accurate, and actionable answers to common breast health questions. The information was rated highly by clinician reviewers and often outperformed search engine results for comprehensiveness and organization. This supports the potential for LLM-powered chatbots or question-answering systems to enhance patient education and engagement. However, limitations were noted regarding outdated content, fabricated references, and inconsistency across similar prompts. There were also concerns about the inability to personalize recommendations and the potential for perpetuating bias. Careful human curation and oversight will be essential to ensure reliability and transparency of LLM-generated patient materials. [25, 26]

More broadly, LLMs pose several challenges that must be addressed before widespread clinical implementation. First of all, the “black box” nature of LLMs, where the reasoning behind outputs is opaque to users. [29] This lack of explainability hinders the ability to audit decisions and identify errors. Techniques to improve LLM interpretability, such as extracting rules or decision trees, are an active area of research. [30] Another challenge is the potential for LLMs to perpetuate biases present in training data, such as over-or under-diagnosis in certain demographics. [31] Careful auditing and debiasing of datasets and models will be required to ensure equitable performance. Data privacy and consent in the use of patient information for model development should also be considered on ethical grounds. [32]

Importantly, LLMs are not static tools, they are evolving technologies undergoing rapid development and refinement. The most recent versions, such as ChatGPT-4, have shown improved performance and safety compared to earlier iterations. Limitations identified in the studies to date may be addressed in future model updates. Additionally, ongoing efforts to build domain-specific LLMs for oncology, such as OncoGPT, [33] may yield stronger results than general-purpose LLMs. However, the fast pace of development also challenges the ability to rigorously validate models before deployment. Prospective studies with standardized evaluation frameworks and reporting will be crucial to establish the utility and safety of LLMs in real-world clinical settings.

This review has several strengths and limitations. We performed a comprehensive search across multiple databases to capture the most recent studies on a rapidly evolving technology. However, relevant studies may have been missed due to inconsistent terminology and lack of established MeSH terms for LLMs. The included studies were highly heterogeneous in LLM type, training data, clinical application, and evaluation metrics. Assessment of study quality and risk of bias was also challenging due to the lack of established tools suited for AI-focused studies. Publication bias is likely present, as studies with positive results may be more likely to be published than negative ones.

## 5. Conclusion

In conclusion, large language models demonstrate promising potential for enhancing diagnosis, management, and patient communication for breast conditions. Across 17 studies, LLMs achieved high diagnostic accuracy, moderate management concordance, and generated high-quality patient information. However, significant risks and limitations were identified, such as biased or fraudulent recommendations, lack of transparency, and insufficient validation in prospective settings. Future work should establish standardized evaluation frameworks and reporting guidelines for LLM studies, curate large diverse training datasets reflective of practice, and incorporate human oversight to mitigate potential harms. With responsible development and validation, LLMs may become powerful tools to support clinicians and patients in the future of breast care.

## Data Availability

All data produced in the present work are contained in the manuscript

## Ethical approval

ethnical approval not required for the study

## Assistance with the study

none

## Presentation

none

